# Identification and prediction of ALS subgroups using machine learning

**DOI:** 10.1101/2021.04.02.21254844

**Authors:** Faraz Faghri, Fabian Brunn, Anant Dadu, PARALS, ERRALS, Elisabetta Zucchi, Ilaria Martinelli, Letizia Mazzini, Rosario Vasta, Antonio Canosa, Cristina Moglia, Andrea Calvo, Michael A. Nalls, Roy H. Campbell, Jessica Mandrioli, Bryan J. Traynor, Adriano Chiò

**Affiliations:** Neuromuscular Diseases Research Section, Laboratory of Neurogenetics, National Institute on Aging, Bethesda, MD 20892, USA; Center for Alzheimer’s and Related Dementias, National Institute on Aging, Bethesda, MD, 20892, USA; Data Tecnica International, Glen Echo, MD, 20812, USA; Department of Computer Science, University of Illinois at Urbana–Champaign, Champaign, IL 61801, USA; Department of Biomedical, Metabolic and Neural Sciences, University of Modena and Reggio Emilia, 41124 Modena, Italy; Neurology Unit, Department of Neurosciences, Azienda Ospedaliero Universitaria di Modena, Modena 41125, Italy; ALS Center, Department of Neurology, Maggiore della Carità University Hospital, Novara 28100 Italy; ‘Rita Levi Montalcini’ Department of Neuroscience, University of Turin, Turin 10126, Italy; Department of Neurology, Johns Hopkins University Medical Center, Baltimore, MD 21287, USA; Reta Lila Weston Institute, UCL Queen Square Institute of Neurology, University College London, London WC1N 1PJ, UK; Institute of Cognitive Sciences and Technologies, C.N.R., Rome 00185, Italy; Neurology 1 and ALS Center, Azienda Ospedaliero Universitaria Città della Salute e della Scienza, Turin 10126, Italy

## Abstract

**Background:** The disease entity known as amyotrophic lateral sclerosis (ALS) is now known to represent a collection of overlapping syndromes. A better understanding of this heterogeneity and the ability to distinguish ALS subtypes would improve the clinical care of patients and enhance our understanding of the disease. Subtype profiles could be incorporated into the clinical trial design to improve our ability to detect a therapeutic effect. A variety of classification systems have been proposed over the years based on empirical observations, but it is unclear to what extent they genuinely reflect ALS population substructure.

**Methods:** We applied machine learning algorithms to a prospective, population-based cohort consisting of 2,858 Italian patients diagnosed with ALS for whom detailed clinical phenotype data were available. We replicated our findings in an independent population-based cohort of 1,097 Italian ALS patients.

**Findings:** We found that semi-supervised machine learning based on UMAP applied to the output of a multi-layered perceptron neural network produced the optimum clustering of the ALS patients in the discovery cohort. These clusters roughly corresponded to the six clinical subtypes defined by the Chiò classification system (bulbar ALS, respiratory ALS, flail arm ALS, classical ALS, pyramidal ALS, and flail leg ALS). The same clusters were identified in the replication cohort. A supervised learning approach based on ensemble learning identified twelve clinical parameters that predicted ALS clinical subtype with high accuracy (area under the curve = 0·94).

**Interpretation:** Our data-driven study provides insight into the ALS population’s substructure and demonstrates that the Chiò classification system robustly identifies ALS subtypes. We provide an interactive website (https://share.streamlit.io/anant-dadu/machinelearningforals/main) so that clinical researchers can predict the clinical subtype of an ALS patient based on a small number of clinical parameters.

**Funding:** National Institute on Aging and the Italian Ministry of Health.

**RESEARCH IN CONTEXT:** *Evidence before this study:* We searched PubMed for articles published in English from database inception until January 5, 2021, about the use of machine learning and the identification of clinical subtypes within the amyotrophic lateral sclerosis (ALS) population, using the search terms “machine learning”, AND “classification”, AND “amyotrophic lateral sclerosis”. This inquiry identified twenty-nine studies. Most previous studies used machine learning to diagnose ALS (based on gait, imaging, electromyography, gene expression, proteomic, and metabolomic data) or improve brain-computer interfaces. One study used machine learning algorithms to stratify ALS postmortem cortex samples into molecular subtypes based on transcriptome data. Kueffner and colleagues crowdsourced the development of machine learning algorithms to approximately thirty teams to obtain a consensus in an attempt to identify ALS patients subpopulation. In addition to clinical trial information in the PRO-ACT database (www.ALSdatabase.org), this effort used data from the Piedmont and Valle d’Aosta Registry for ALS (PARALS). Four ALS patient categories were identified: slow progressing, fast progressing, early stage, and late stage. This approach’s clinical relevance was unclear, as all ALS patients will necessarily pass through an early and late stage of the disease. Furthermore, no attempt was made to discern which of the existing clinical classification systems, such as the El Escorial criteria, the Chiò classification system, and the King’s clinical staging system, can identify ALS subtypes. We concluded that there remained an unmet need to identify the ALS population’s substructure in a data-driven, non-empirical manner. Building on this, there was a need for a tool that reliably predicts the clinical subtype of an ALS patient. This knowledge would improve our understanding of the clinical heterogeneity associated with this fatal neurodegenerative disease.

*Added value of this study:* This study developed a machine learning algorithm to detect ALS patients’ clinical subtypes using clinical data collected from the 2,858 Italian ALS patients in PARALS. Ascertainment of these patients within the catchment area was near complete, meaning that the dataset truly represented the ALS population. We replicated our approach using clinical data obtained from an independent cohort of 1,097 Italian ALS patients that had also been collected in a population-based, longitudinal manner. Semi-supervised learning based on Uniform Manifold Approximation and Projection (UMAP) applied to a multilayer perceptron neural network provided the optimum results based on visual inspection. The observed clusters equated to the six clinical subtypes previously defined by the Chiò classification system (bulbar ALS, respiratory ALS, flail arm ALS, classical ALS, pyramidal ALS, and flail leg ALS). Using a small number of clinical parameters, an ensemble learning approach could predict the ALS clinical subtype with high accuracy (area under the curve = 0·94).

*Implications of all available evidence:* Additional validation is required to determine these algorithms’ accuracy and clinical utility in assigning clinical subtypes. Nevertheless, our algorithms offer a broad insight into the clinical heterogeneity of ALS and help to determine the actual subtypes of disease that exist within this fatal neurodegenerative syndrome. The systematic identification of ALS subtypes will improve clinical care and clinical trial design.

## INTRODUCTION

Amyotrophic lateral sclerosis (ALS, OMIM #105400) is one of the most common forms of neurodegeneration in the population, accounting for approximately 6,000 deaths in the United States and 11,000 deaths in Europe annually.^1, 2^ Characterized by progressive paralysis of limb and bulbar musculature, it typically leads to death within three to five years of symptom onset. Medications only minimally slow the rate of progression, and, as a consequence, treatment focuses on symptomatic management.

Genetic advancements have shown that ALS is not a single entity but consists of a collection of syndromes in which the motor neurons degenerate.^3^ Together with these multiple genetic etiologies, there is a broad variability in the disease’s clinical manifestations in terms of the age of symptom onset, site of onset, rate and pattern of progression, and cognitive involvement. This clinical heterogeneity has hampered efforts to understand the cellular mechanisms underlying this fatal neurodegenerative syndrome and has hindered clinical trial efforts to find useful therapies.^4^

Given the importance of this clinical heterogeneity within ALS, it is not surprising that there has been considerable effort to develop classification systems for patients over the years. Examples include grouping based on family status^5^, clinical milestones^6^, neurophysiological measurements^7, 8^, and diagnostic certainty.^9^ Though useful, each of these existing classification systems suffers from a central problem. It is unclear if they identify meaningful subgroups within the ALS population or represent human constructs applied to the data based on empirical observations. The ability to determine the correct number and nature of subgroups within the ALS population would be a significant step toward understanding the disease. By extension, a reliable method to predict an individual patient’s subgroup would be useful for clinical care and clinical trial design.

Here, we explored the clinical patterns of ALS by applying unsupervised and semi-supervised machine learning to deeply-phenotyped, population-based collections of patients (see Figure 1 for the analysis workflow). Our goal was to determine which subtypes of the disease exist within this patient population. The advantage of these machine learning approaches is their ability to identify complex relationships in an unbiased and data-driven manner that moves beyond the traditional univariate approach. Following the successful identification of the ALS subtypes, we used supervised machine learning to build predictor models that can accurately classify individual patients and deployed this as a simple-to-use website that clinicians can access.

**Figure 1.**
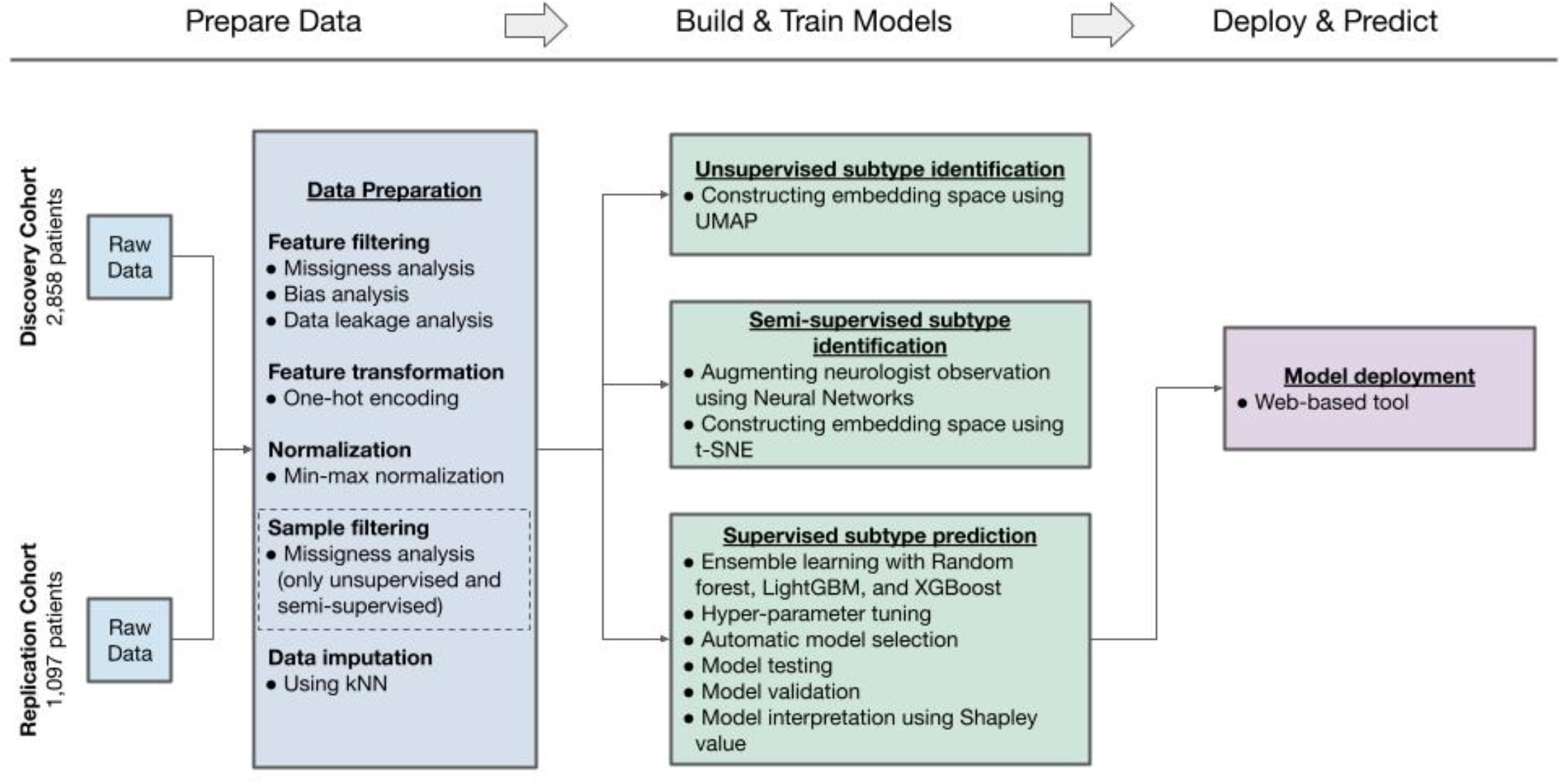
Workflow followed in this study. Unsupervised and semi-supervised machine learning was applied to clinical data collected from two population-based ALS registries to identify clinical subtypes. Supervised machine learning was used to predict subtypes based on clinical parameters, and a web-based tool was built for clinical researchers to apply to their own data.

## METHODS

### Study participants

The discovery cohort consisted of patients who had been diagnosed with ALS and were enrolled in the Piedmont and Valle d’Aosta Registry for ALS (PARALS) between January 1, 1995, and December 31, 2015.^10^ The replication cohort consisted of incident cases who had been diagnosed with ALS and were enrolled in the Emilia Romagna Region ALS (ERRALS) registry between January 1, 2009, and March 1, 2018.^11^ A vital feature of these long-standing, population-based epidemiological studies is their collection of detailed data on patients throughout their illness. Informed written consent was obtained from the participants. The studies were approved by the ethics committees of Azienda Ospedaliera Universitaria City of Health and Science of Turin and Azienda Ospedaliero Universitaria of Modena. All records were anonymized according to the Italian code for the protection of personal data and data were treated following the UE 2016/679 General Data Protection Regulation (GDPR).

### Filtering and pre-processing of the clinical data

The clinical data were filtered before analysis. Features with high missingness (e.g., cancer type), high sampling bias (e.g., place of birth), and features that could introduce data leakage (e.g., tracheostomy, initial diagnosis was primary lateral sclerosis) were omitted from all of the analyses. For unsupervised and semi-supervised subtype identification, samples with missing values in the revised ALS Functional Rating Score (ALSFRS-R^12^) feature were also excluded (n = 497 in the discovery cohort, n = 108 in the replication cohort). Ultimately, 2,361 cases in the discovery cohort and 989 in the replication cohort passed the filtering steps and were available for the identification analysis. In contrast, samples with missing ALSFRS-R data were included in the supervised machine learning analysis as these algorithms are capable of handling missingness. The prediction modeling used 2,858 cases in the discovery cohort and 1,097 in the replication cohort. Categorical features were encoded to numerical using the *one-hot encoding* method. *Min-max normalization* was applied to numeric features to preserve the distribution’s shape and ensure a zero-to-one range.

### Data imputation

After filtering and pre-processing, the following features had residual missingness that was distributed randomly across the patients at a rate of 15–20%: (i) forced vital capacity (FVC) percent at diagnosis, (ii) body mass index (BMI) at two years before illness, (iii) rate of decline of BMI per month, (iv) weight two years before illness, (v) BMI at diagnosis, (vi) height, and (vii) weight at diagnosis. We used the *k-Nearest Neighbor* (kNN) imputation method with k = 5 neighbors to preserve the clusters.^13^

### Unsupervised machine learning

The data analysis pipeline for this work was performed in Python 3.6 with the support of several open-source libraries (numpy, pandas, matplotlib, seaborn, plotly, scikit-learn, umap, xgboost, lightgbm, and tensorflow). To facilitate replication and expansion of our work, we have made the notebook publicly available under the GPLv3 license on Google Colaboratory and GitHub at https://github.com/ffaghri1/ALS-ML. This code includes the rendered Jupyter notebook with a full step-by-step description of the data pre-processing, statistical, and machine learning analysis used in this study. The machine learning parameters are described in the Python Jupyter notebook. Manuscript visualizations were created with tidyverse (version 1.3), ggplot2 (version 3.3.2), and plotly (version 4.9.2.2) as implemented in R (version 4.0.3).

### Semi-supervised machine learning

Data were initially processed using a *multilayer perceptron neural network* consisting of five hidden layers with 200, 100, 50, 25, and 3 neurons (Supplemental Figure S1).^14^ This supervised technique reduced the sixty-six clinical features of the ALS datasets down to three dimensions. The dimension-reduction network was trained on the ‘*clinical type at one-year*’ feature using a *Softmax* classifier. After training the model with ten-fold cross-validation, the data was used as input for the network’s forward loop. The last hidden layer activations, which represents the reduction to three dimensions, were used as input for the UMAP algorithm.^15^

### Supervised subtype prediction

Ensemble learning was used to develop predictive models forecasting the ALS clinical subtype of a patient based solely on the clinical data obtained at the first neurology visit. Ensemble learning combines multiple learning algorithms to generate a better predictive model than could be obtained using a single learning algorithm.^16^ To do this, the stacking ensembles of three supervised machine learning algorithms (Random forest^17^, LightGBM^18^, and XGBoost^19^) were evaluated, and the best performing ensemble model was selected. Internal and external validation was used to assess performance and determine the best algorithms and parameters to use in the model (see **Supplementary Methods** and Supplemental Figure S2).

The Shapley additive explanations (SHAP) approach was used to evaluate each feature’s influence in the ensemble learning. This approach, used in game theory, assigned an importance (Shapley) value to each feature to determine a player’s contribution to success.^20^ Shapley explanations enhance understanding by creating accurate explanations for each observation in a dataset. They bolster trust when the critical variables for specific records conform to human domain knowledge and reasonable expectations. The interactive website (https://share.streamlit.io/anant-dadu/machinelearningforals/main) was developed as an open-access and cloud-based platform.

## RESULTS

We aimed to identify the clinical subtypes that exist within the ALS patient population in a data-driven manner. To do this, we applied unsupervised and semi-supervised machine learning approaches to a cohort consisting of 2,858 patients diagnosed with ALS and enrolled in the Piedmont and Valle d’Aosta Registry for ALS (PARALS) over twenty five years. This registry is a population-based epidemiological study established in 1995 to prospectively ascertain ALS cases within Northwestern Italy (including the cities of Turin and Aosta). The registry has near-complete case ascertainment within its catchment population of nearly 4·5 million inhabitants.^10^ Patients are followed throughout their illness, allowing for longitudinal data collection. Supplementary Table S1 summarizes the clinical and demographic details of the discovery cohort. The sixty-six clinical features collected for each case are listed in Supplemental Table S2, and an exploratory data analysis describing the content of each feature is provided in the **Supplemental Material**.

The unsupervised approach used a UMAP algorithm to visualize high-dimensional data within a lower-dimensional topological space.^15^ This nonlinear dimension reduction preserves the local and global structures existing within the data to yield reproducible and meaningful clusters. The semi-supervised approach involved pre-processing the data with a *multi-layered perceptron neural network*^15^ and using the dimension-reduced output of this neural network as the UMAP algorithm’s input. This technique, motivated by the Augmented Intelligence in Healthcare ethos, leverages the clinician’s diagnostic acumen to improve the model.^21, 22^

The primary outcome measure of our analysis was a comparison of the subtype clusters defined by the machine learning approaches to the six clinical subtypes (bulbar ALS, respiratory ALS, flail arm ALS, classical ALS, pyramidal ALS, and flail leg ALS) assigned manually by neurologists according to the Chiò classification system.^23^ Unlike other classification systems, these subtypes are determined based on observing the patient’s illness over the first year after symptom onset. The clinical subtypes assigned by the Chiò classification system were not entered into the unsupervised and semi-supervised algorithms and were not used to construct the patient clusters. Instead, the clinical subtypes were used to color the dots representing the patients produced by the machine learning algorithms.

Both the unsupervised and semi-supervised approaches identified multiple clusters of patients, representing distinct subtypes of ALS (see Figure 2A for the neural network-UMAP and Supplemental Figure S3 for the results of the UMAP alone). Color-coding the ALS patients according to the clinical subtype assigned by the patient’s neurologist showed that the clusters roughly corresponded to the six clinical subtypes previously defined by the Chiò classification system. Visually investigating these three-dimensional projections, the optimum separation of the ALS patients into their clinical subtypes was obtained using the semi-supervised machine learning approach. There was excellent discrimination of the bulbar ALS, respiratory ALS, flail arm ALS, and classical ALS subtypes. In contrast, the pyramidal ALS and flail leg ALS overlapped significantly, though the flail leg ALS variant did form a distinct tail that did not overlap with the other subtypes. Overall, we found that 787 (99·7%) of the bulbar ALS cases, 42 (100·0%) of the respiratory cases, 150 (91·5%) of the flail arm cases, and 663 (93·4%) of the classical cases were assigned to the same subtype by the clinician and the semi-supervised algorithm.

**Figure 2.**
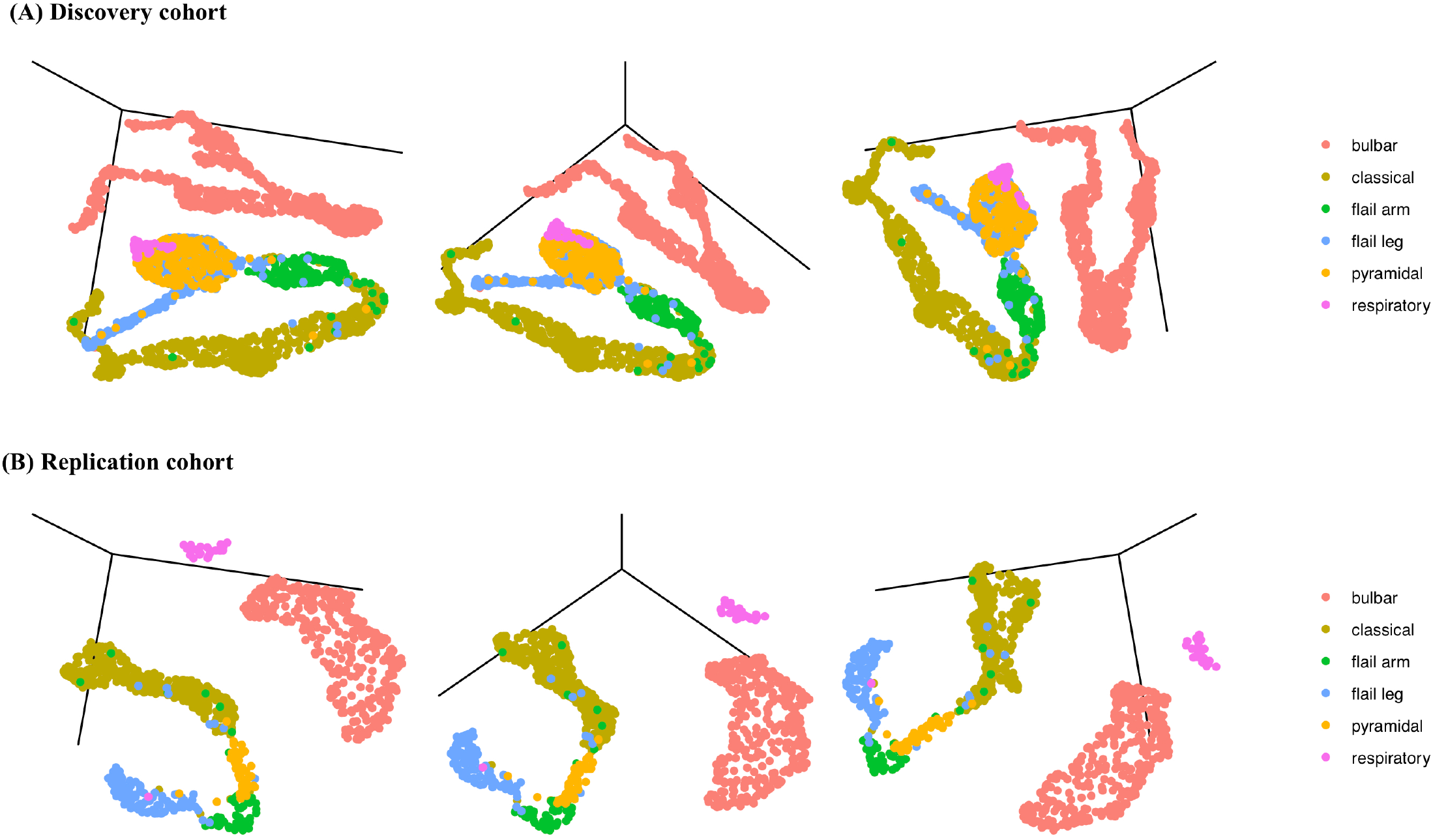
ALS subtypes identified by machine learning in the discovery and replication cohorts. The top row (A) shows the three different 3D projections of the discovery cohort defined by the semi-supervised machine learning algorithm consisting of a UMAP algorithm applied to the output of a five-layer neural network. The same 3D projections of the replication cohort are shown in the bottom row (B). Each patient (dot) was color-coded after machine learning cluster generation according to the Chiò classification system. Interactive three-dimensional graphs are available on https://share.streamlit.io/anant-dadu/machinelearningforals/main.

To validate our results, we replicated the ALS subtype identification in an independent cohort consisting of 1,097 incident ALS cases gathered over nine years by a second prospective, population-based ALS registry based in the Emilia Romagna Region (ERRALS). This registry prospectively collects demographic and clinical data on ALS incident cases within a Northwestern region of Italy that includes the cities of Modena and Bologna.^11^ Like PARALS, the catchment population of ERRALS is 4·4 million, and patients are followed longitudinally throughout their illness. The methods for collecting clinical and demographic data were standardized across the registries to facilitate comparisons between these two epidemiological efforts. Figure 2B shows the subtypes and clusters identified in the independent replication cohort. The cluster pattern is similar to that observed in the discovery cohort, confirming the reproducibility of our data-driven approach. Interactive three-dimensional graphs are available on https://share.streamlit.io/anant-dadu/machinelearningforals/main (see “Explore the ALS subtype topological space).

Our semi-supervised machine learning algorithm was more accurate than the other dimension reduction approaches such as principal component analysis (PCA) and independent component analysis (ICA) (Supplemental Figure S4). Furthermore, other ALS classification schema, such as the El Escorial categories^9^, family status^5^, the presence or absence of the pathogenic *C9orf72* repeat expansion, MiToS clinical staging^24^, ALSFRS-R score^12^, and King’s clinical stages^6^, did not label the clusters in a meaningful, clinically-useful manner (Figure 3).

**Figure 3.**
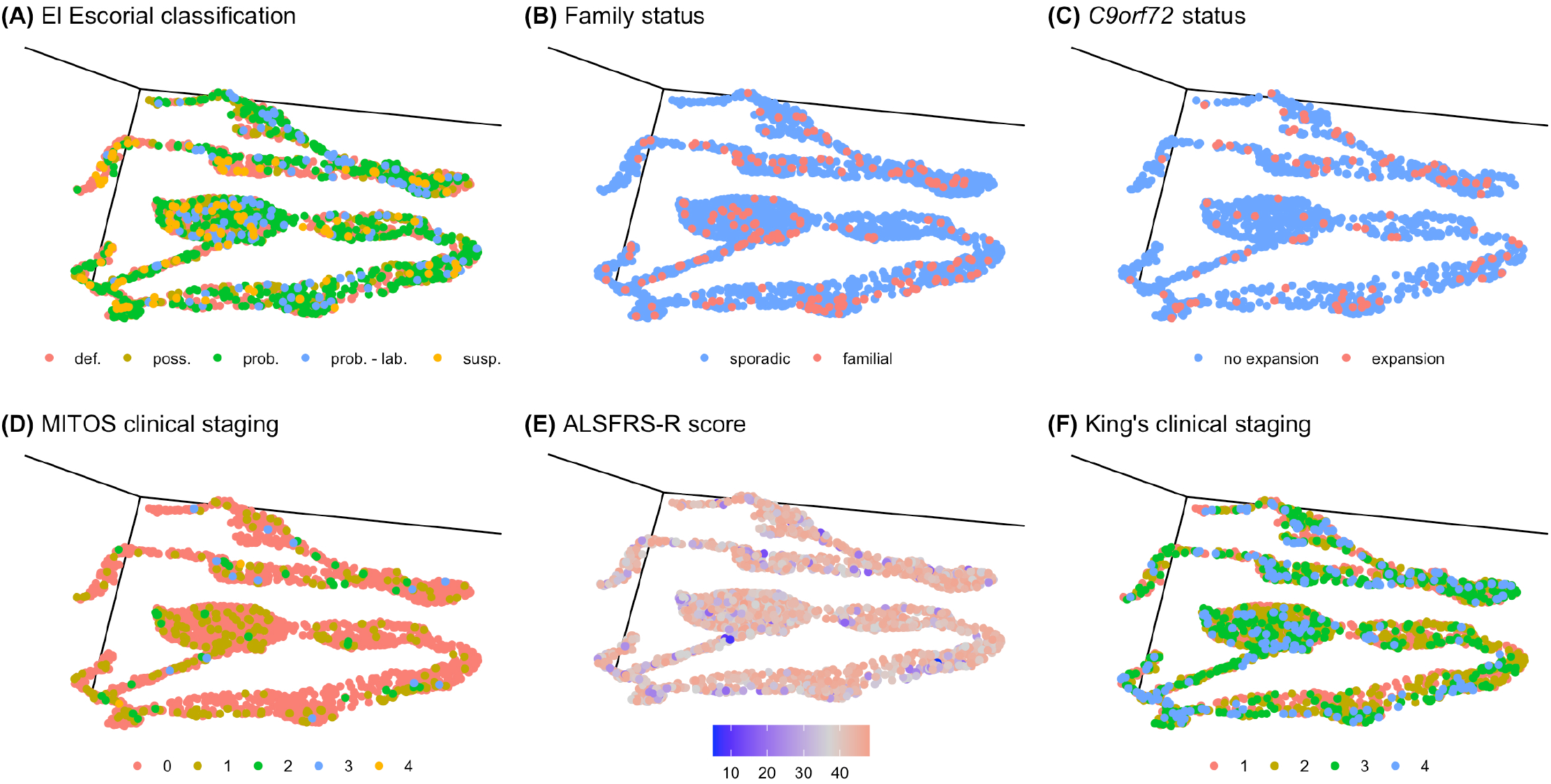
Different classification schema applied to the semi-supervised 3D projection of the ALS discovery cohort. (A) The El Escorial classification system assigns patients to definite (def.), probable (prob.), probable - laboratory supported (prob. - lab.), possible (poss.), and suspected (susp.) categories based on their disability. (B) Patients with a family history of ALS are represented by red dots, and patients with sporadic disease are shown by blue dots. (C) Patients carrying the pathogenic repeat expansion are represented by red dots. (D) The MITOS classification system assigns patients to clinical stages 0 to 4 based on their disability. (E) The ALSFRS-R score rates the severity of disability ranging from 0 to 48 (no disability). (F) The King’s clinical staging system classifies patients into four stages according to their disability level.

Next, we applied a supervised learning approach called *ensemble learning* to develop predictive models forecasting the ALS clinical subtype of a patient based solely on the clinical data obtained at the first neurology visit. Ensemble learning combines multiple learning algorithms (Random forest^17^, LightGBM^18^, and XGBoost^19^) to generate a better predictive model than obtained using a single learning algorithm.^16^ When all available features (n = 66) are included in the model, the clinical subtype of a patient is predicted with high accuracy (internal validation area under the curve (AUC) = 0·98, external validation AUC = 0·94, and see Supplemental Figure S5 for the Receiver Operator Characteristic (ROC) curves).

To increase this approach’s clinical utility, we decreased the number of parameters included in the model without sacrificing accuracy using a randomized grid search and a game theory approach based on Shapley analysis.^20^ The predictor model built with the top eleven factors was equally robust compared to the all-inclusive model (internal validation AUC = 0·98 and external validation AUC = 0·95, Figure 4 and Supplemental Figure S6). Table 1 and Figure 5 lists the eleven parameters selected for the final model and their relative contributions to the model’s precision. Finally, we implemented an interactive website (https://share.streamlit.io/anant-dadu/machinelearningforals/main, see “Predict Patient ALS Subtype”) that allows clinical researchers to determine an ALS patient’s future clinical subtype based on these eleven parameters. We have also developed a “what-if analysis” functionality, helping to better understand the effects of changes on the outcome.

**Table 1.** Clinical features selected for the final model with their relative contributions to the model’s precision.

|  | **Model precision** |  |
| --- | --- | --- |
|  | **Relative Importance** | **Standard Deviation of Relative Importance** |
| **Anatomical level at onset** | 1·0 | 0·07801415 |
| **Site of symptom onset** | 0·4596638 | 0·02144825 |
| **Onset side** | 0·1322353 | 0·02321156 |
| **Weight at diagnosis (kg)** | 0·04158558 | 0·0004548315 |
| **El Escorial category at diagnosis** | 0·03313919 | 0·002519089 |
| **ALSFRS-R part 1 score** | 0·02724375 | 0·0008967141 |
| **Time of first ALSFRS-R measurement**<br>**(days from symptom onset)** | 0·01998734 | 0·0 |
| **Smoking status** | 0·01920519 | 0·0001980213 |
| **Age at symptom onset** | 0·01484295 | 0·0 |
| **Rate of BMI decline (per month)** | 0·01409138 | 0·0 |
| **FVC% at diagnosis** | 0·01347821 | 0·0 |

ALSFRS-R part 1 score refers to the first question in the ALS Functional Rating Scale – Revised rating scale concerning speech.

**Figure 4.**
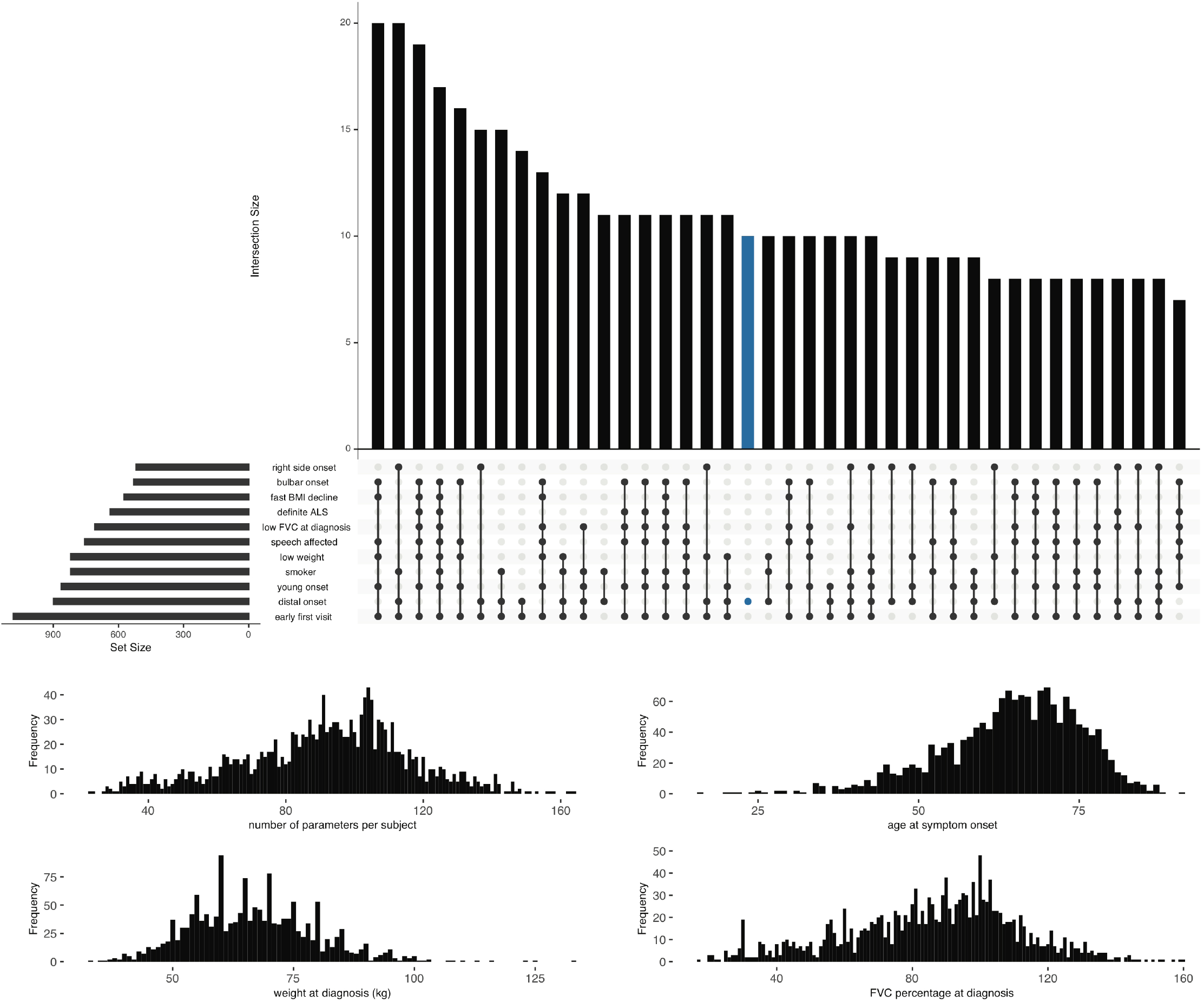
Clinical parameters used in the supervised machine learning model to predict ALS clinical subtype. (A) Graphical representation of the overlap between the eleven parameters with the most significant impact on the classification model. The dark circles in the dot plot indicate the parameters that are part of an intersection, and the vertical bar plot reports the number of patients with that parameter combination. The horizontal bar plot reports the set sizes. Analysis was confined to 699 ALS patients with no missing data. (B) Distribution of the parameters in each patient. On average, a patient had five of these clinical features. (C - E) Distribution of the age at onset, weight at diagnosis, and forced vital capacity percent at diagnosis in the analyzed patients.

**Figure 5.**
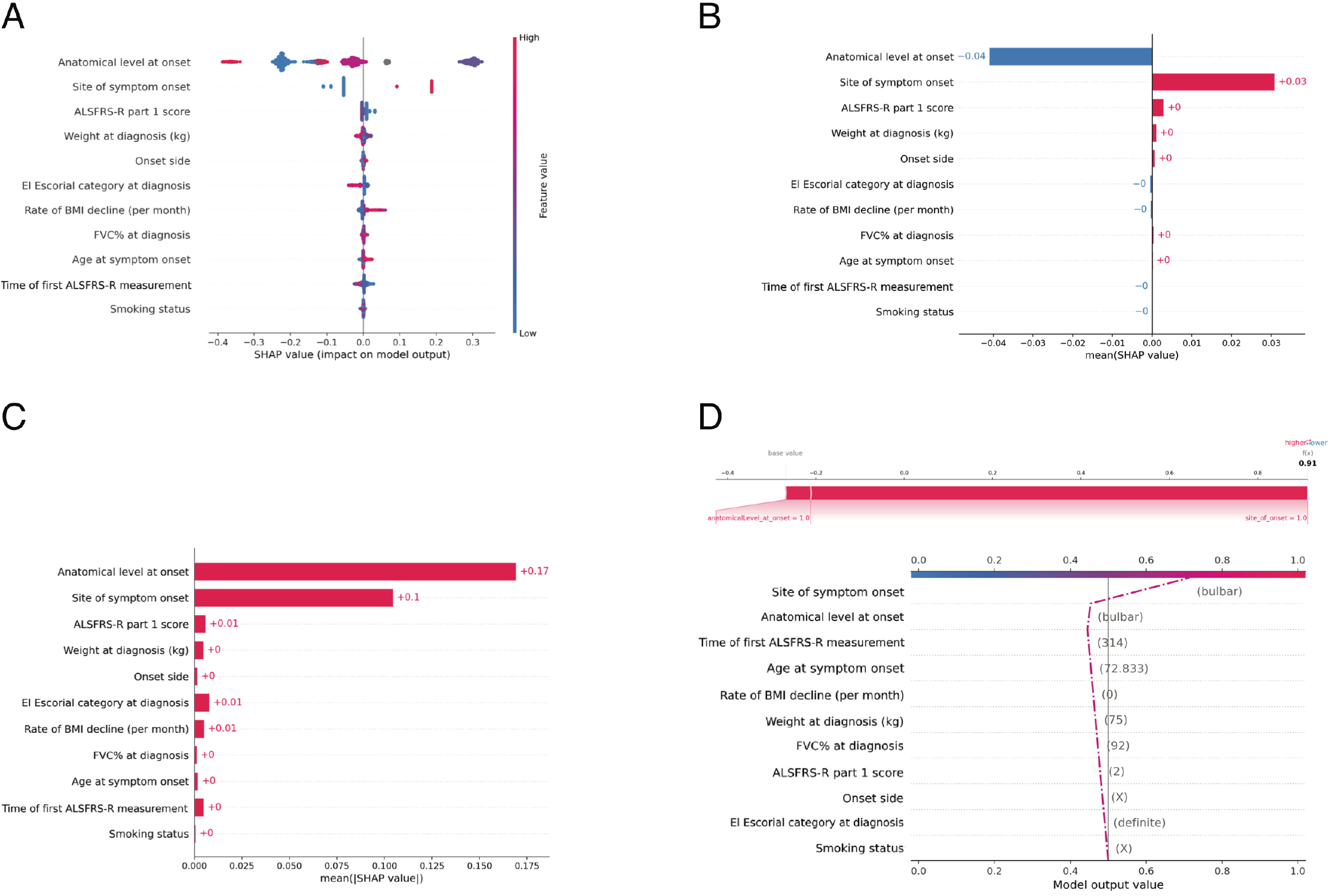
Eleven features used in the supervised machine learning model to predict ALS clinical subtype. (A) Distribution of the Shap values for the eleven features with the most significant impact on the classification model. Each point represents a subject and may have a positive or negative impact depending on its SHAP value. For instance, high values of the rate of BMI decline in red contribute strongly to the positive class, while low values in blue contribute to a lesser extent to the negative class. (B & C) The aggregate of the Shap values is shown for the top eleven features (ranked from most to least important). (D) Model output trajectory for a single subject with the bulbar subtype of ALS. The predicted probability that the patient had the bulbar subtype of ALS was 0.91, predominantly driven by the patient’s bulbar site of symptom onset and only minorly driven by their smoking status and El Escorial category at diagnosis. See https://share.streamlit.io/anant-dadu/machinelearningforals/main for more examples.

## DISCUSSION

The ALS community has long sought a reliable method to identify the subgroups existing within the ALS population. Knowledge of the substructure of ALS would improve our understanding of the clinical heterogeneity associated with this fatal neurodegenerative disease. By extension, it would enhance patient care and provide insights into the underlying pathological mechanisms.^25, 26^ Here, we used a machine learning approach to identify such subtypes within a large cohort of ALS patients, and we then replicated our findings in an independent, population-based cohort. This unbiased, data-driven approach confirmed the existence of subtypes within the ALS spectrum of disease. Interestingly, these subtypes roughly corresponded to those previously defined by the Chiò classification system^23^, demonstrating that schema’s utility. Unlike other subtyping approaches, the Chiò classification system relies on the patient’s clinical data collected during the first year of illness.^23^ This one-year observation period allows the disease’s symptoms to manifest more clearly and provides an accurate assessment of the progression rate. Though progression is a fundamental feature of ALS, it is not typically employed in determining the subtype of the disease.

The primary obstacles to deciphering the clinical heterogeneity observed among patients with ALS have been the lack of a sufficiently large dataset and the inability to analyze multi-dimensional relationships within that data. We used data from two large, population-based registries that had enrolled ALS patients over ten years to address these issues. These registries prioritized data collection throughout the patient’s illness, and overall, they contained nearly 300,000 pieces of information that were central to our categorization efforts. Our results highlight the value of disease registries that capture deep phenotype across an entire catchment area. Previous efforts to catalog the various subgroups of ALS hinged on a small number of clinical features, such as family history or site of symptom onset.^5–9^ Although clinically useful, these univariate or oligovariate classification systems do not capture the complicated clinical patterns within the ALS population. In contrast, the new machine learning algorithms that we applied are adept at deciphering complex and multi-faceted relationships.

Remarkably, our machine learning algorithm defined the same subgroups outlined by Chiò and colleagues. We do not maintain that our machine learning approach is better at identifying categories than this group of experienced ALS neurologists. Instead, we validated the Chiò classification system^23^ using an unbiased, data-driven approach and provide *prima facie* evidence that this schema captures the ALS population’s substructure. Classification based on other schemes, such as the El Escorial, MiToS, and King’s systems, was not useful in assigning patients to a disease subtype (see Figure 3). Nevertheless, our machine learning algorithm provides opportunities to improve and refine the Chiò classification system, especially as the pyramidal and the flail leg subtypes may not be distinct from each other as other subtypes. This finding was unexpected as these patients are easily distinguished from each other in the clinic, highlighting machine learning’s ability to provide new and essential insights into a complex disease. It also offers a novel starting point for exploring the pyramidal and flail leg ALS variants’ underlying neurobiology.

Having established that the six subtypes outlined by the Chiò classification reflect the correct substructure of ALS, we next considered how clinicians and researchers could use this information. The ability to assign patients to subgroups helps unravel the disease’s clinical heterogeneity and will aid in discussions with affected individuals about likely disease course and prognosis. For example, patients with the respiratory subtype of ALS had a faster rate of progression. They were more likely to require non-invasive positive pressure ventilation (NIPPV) and gastrostomy feeding at an earlier stage than ALS cases with the upper motor predominant form of the disease. Outcome data from previously negative clinical trials can be reanalyzed to look for a therapeutic effect limited to one or two subgroups. A similar approach has been successful in Parkinson’s disease.^27^ Genetic heterogeneity also handicaps our ability to implicate new loci in the disease’s pathogenesis using genome-wide association analysis. Including the subgroup as a covariate or restricting the search to a single subtype may resolve this issue as it leads to a more homogeneous patient population.

It has not escaped our attention that the topology representation of the ALS subtypes produced by the machine learning algorithm resembles the central nervous system (CNS). We observed this pattern most clearly in Figure 2. The bulbar subtype delineates the cerebrum, and the spinal cord is represented by a long tail running successively from flail arm, pyramidal, classical, to flail leg subtypes. We speculate that this arrangement hints at a broader anatomical organization within the ALS spectrum, perhaps reflecting subtle differences of the motor neuron subtypes within each segment of the CNS and differing susceptibilities to pathogenic mechanisms.

Our study has several limitations. Machine learning algorithms can identify patterns within a dataset even when no such pattern exists. Such ‘overfitting of the model’ is an inherent problem with this statistical methodology, and the most legitimate remedy is to attempt replication in an independent dataset. Our study’s strength was the availability of such data from a second, population-based ALS registry, and analysis of this independent cohort yielded remarkably similar outcomes to the discovery cohort, demonstrating the robustness of our approach. In addition, the handling of missing data is increasingly recognized as a critical constraint of machine learning. Our data was remarkably complete, as shown in the exploratory data analysis notebooks. Nonetheless, as with any real-life clinical dataset, information was missing for some parameters. We were transparent and cautious in handling these issues, and we followed the recommendations recently published by the American Heath Association.^28^

Our machine learning used the same set of patients initially used by Chiò and colleagues to define their subtypes.^23^ It is unlikely that this led to recruitment bias as information concerning the Chiò subtypes was not used to generate our models, and the clinical parameters used to create the models are standard across the ALS field. Furthermore, population-based registries decrease the possibility of recruitment bias as they capture every case within a catchment area. Nevertheless, both our discovery and replication data originated from the Northern Italian population, and additional studies in other countries are required to test our approach’s generalizability. These data will likely have to be collected prospectively, as there is generally insufficient information to determine the Chiò classification of samples in retrospective databases such as PRO-ACT (https://nctu.partners.org/proact).^29^

Like other statistical systems, machine learning algorithms are only practical if they can be applied broadly. To facilitate this, we have established a website where a neurologist can enter a patient’s characteristics to predict their subtype membership. We have made our programming code publicly available (https://github.com/ffaghri1/ALS-ML) so that other researchers can apply it and modify it as our understanding of ALS and the application of machine learning evolves. Though our current categorization approach is robust, we anticipate that it will improve over time to the point that it will become an indispensable tool for clinicians dealing with ALS patients. Here, we provide an early demonstration of machine learning to unravel highly complex and interrelated disease systems such as ALS.

## Data Availability

To facilitate replication and expansion of our work, we have made the notebook publicly available under the GPLv3 license on Google Colaboratory and GitHub at https://github.com/ffaghri1/ALS-ML. This code includes the rendered Jupyter notebook with a full step-by-step description of the data pre-processing, statistical, and machine learning analysis used in this study. We have also developed an interactive website ((https://share.streamlit.io/anant-dadu/machinelearningforals/main) as open access and cloud-based platform that allows clinical researchers to determine an ALS patient's future clinical subtype based on a small number of clinical parameters.

https://share.streamlit.io/anant-dadu/machinelearningforals/main

## CONTRIBUTORS

AC and BJT designed and oversaw the study. FF, FB, MAN, RHC, JM, BJT and AC performed the primary interpretation of the data. FF and BJT wrote the manuscript. AC and JM made major contributions to manuscript editing. EZ, IM, LM, RV, AC, CM, AC, JM, and AC recruited and phenotyped the participants. All authors contributed to and critically reviewed the final version of the manuscript.

## CONSORTIUM

The members of the PARALS Consortium are Adriano Chiò, Andrea Calvo, Cristina Moglia, Antonio Canosa, Umberto Manera, Rosario Vasta, Alessandro Bombaci, Maurizio Grassano, Maura Brunetti, Federico Casale, Giuseppe Fuda, Paolina Salamone, Barbara Iazzolino, Laura Peotta, Paolo Cugnasco, Giovanni De Marco, Maria Claudia Torrieri, Francesca Palumbo, Salvatore Gallone, Marco Barberis, Luca Sbaiz, Salvatore Gentile, Alessandro Mauro, Letizia Mazzini, Fabiola De Marchi, Lucia Corrado, Sandra D’Alfonso, Antonio Bertolotto, Maurizio Gionco, Daniela Leotta, Enrico Oddenino, Daniele Imperiale, Roberto Cavallo, Pietro Pignatta, Marco De Mattei, Claudio Geda, Diego Maria Papurello, Salvatore Amarù, Graziano Gusmaroli, Cristoforo Comi, Carmelo Labate, Fabio Poglio, Luigi Ruiz, Delfina Ferrandi, Lucia Testa, Eugenia Rota, Marco Aguggia, Nicoletta Di Vito, Piero Meineri, Paolo Ghiglione, Nicola Launaro, Michele Dotta, Alessia Di Sapio, Guido Giardini, Patrizia Julita, and Claudio Solaro.

The members of the ERRALS Consortium are: Jessica Mandrioli, Nicola Fini, Ilaria Martinelli, Elisabetta Zucchi, Giulia Gianferrari, Cecilia Simonini, Stefano Meletti, Rocco Liguori, Veria Vacchiano, Fabrizio Salvi, Ilaria Bartolomei, Roberto Michelucci, Pietro Cortelli, Rita Rinaldi, Anna Maria Borghi, Andrea Zini, Elisabetta Sette, Valeria Tugnoli, Maura Pugliatti, Elena Canali, Luca Codeluppi, Franco Valzania, Lucia Zinno, Giovanni Pavesi, Doriana Medici, Giovanna Pilurzi, Emilio Terlizzi, Donata Guidetti, Silvia De Pasqua, Mario Santangelo, Patrizia De Massis, Martina Bracaglia, Mario Casmiro, Pietro Querzani, Simonetta Morresi, Marco Longoni, Alberto Patuelli, Susanna Malagù, Marco Currò Dossi, Simone Vidale, and Salvatore Ferro.

## ACKNOWLEDGEMENTS

This work was supported in part by the Intramural Research Programs of the NIH, National Institute on Aging (Z01-AG000949-02). This work was in part supported by the Italian Ministry of Health (Ministero della Salute, Ricerca Sanitaria Finalizzata, grant RF-2016-02362405), the European Commission’s Health Seventh Framework Programme (FP7/2007–2013 under grant agreement 259867), and the Joint Programme - Neurodegenerative Disease Research (Strength, ALS-Care and Brain-Mend projects), granted by Italian Ministry of Education, University and Research. This study was performed under the Department of Excellence grant of the Italian Ministry of Education, University and Research to the ‘Rita Levi Montalcini’ Department of Neuroscience, University of Torino, Italy. The Emilia Romagna Registry for ALS (ERRALS) is supported by a Grant from the Emilia Romagna Regional Health Authority. This study was also partly funded by the AGING Project for Department of Excellence at the Department of Translational Medicine (DIMET), Università del Piemonte Orientale, Novara, Italy. This study used the high-performance computational capabilities of the Biowulf Linux cluster at the National Institutes of Health, Bethesda, Maryland, USA (http://biowulf.nih.gov). We thank the Laboratory of Neurogenetics (NIH) staff for their collegial support and technical assistance.

## Supplemental data

### Supplemental Methods

#### Supervised subtype prediction

Internal and external validation was used to assess performance and to determine the best algorithms and parameters to use in the model. The internal validation was also combined with random hyperparameter tuning, and feature selection and generation as follows:

a. The effects of the different algorithms, algorithm parameters, and features were evaluated. In multiple iterations, the cohort data was split into training and validation datasets.
b. Hyperparameters were tuned on the training cohort using three-fold cross-validation. Two folds were used for the optimization, and the third was used to validate the results. To tune the hyperparameters, a grid of possible parameter combinations was built, and a randomized grid search was used to find the combination that maximized accuracy.
c. The features included in each iteration were updated using the variable importance obtained from the previous iteration as a probabilistic prior. The best performing model and features are then passed to the feature evolution stage. A genetic algorithm was used in the feature evolution stage to find the best set of model parameters and feature transformations for the final model. These genetic algorithms are a search heuristic inspired by Charles Darwin’s theory of natural evolution applied to non-genetic data.
d. For the final external validation, the model was trained using the complete discovery dataset and the optimal hyperparameter combination. The resulting model was then evaluated on the replication set.

**Supplemental Figure S1.**
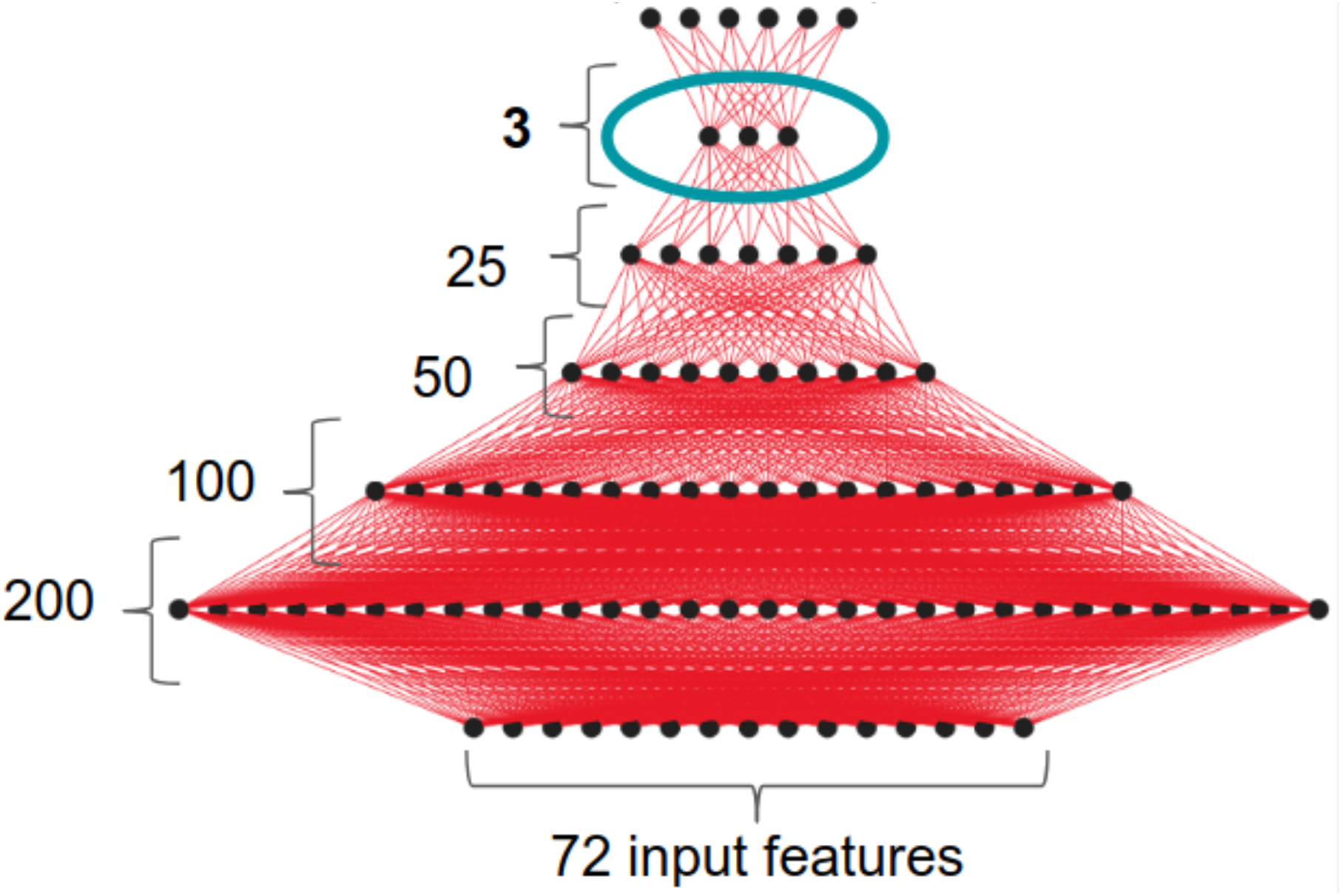
Architecture of the multilayer perceptron neural network used for the semi-supervised subtype identification of ALS. The neural network consists of five hidden layers with 200, 100, 50, 25 and 3 neurons. The neural network acts as a dimension reduction technique, compressing the data to 3 dimensions. After training the network with ten-fold cross validation, the activations of the last hidden layer are used as input for the UMAP algorithm.

**Supplemental Figure S2.**
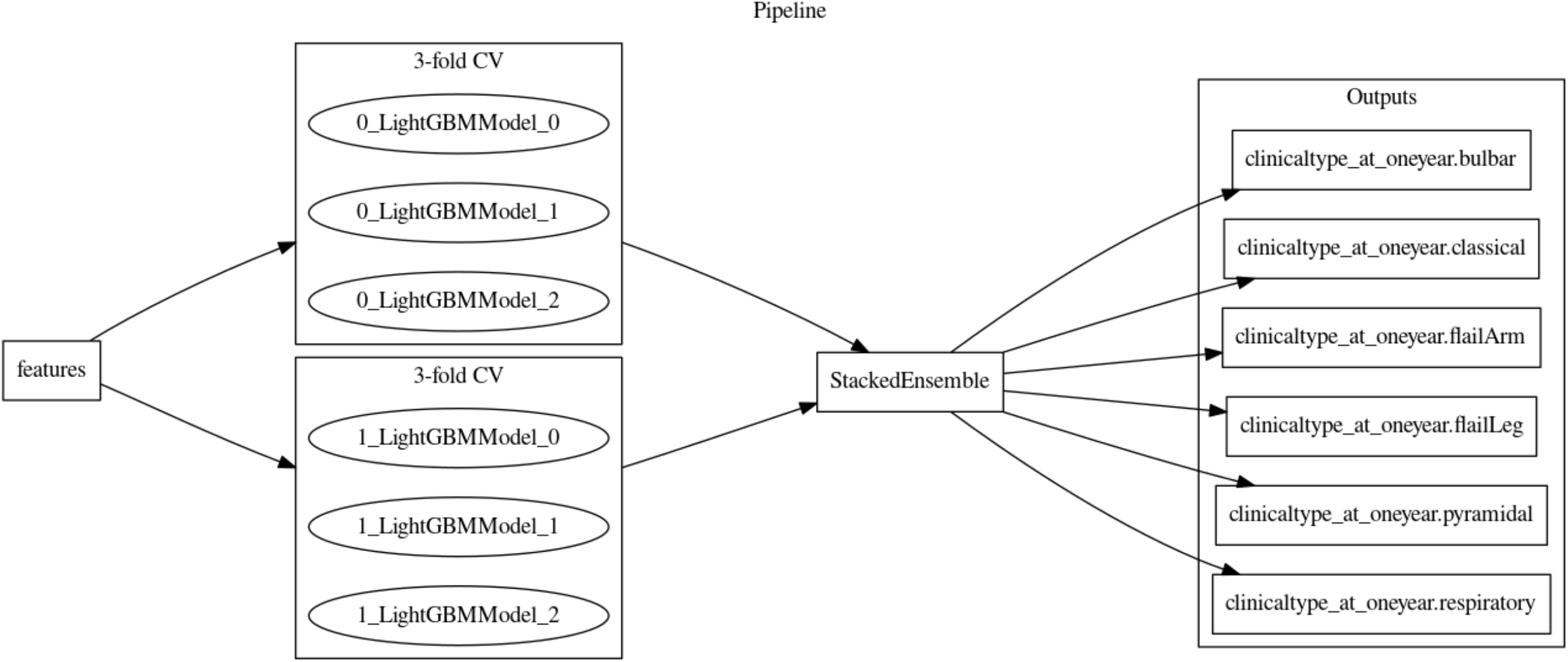
Supervised learning model used for ALS subtype identification. The predictive model using all the clinical features was a stacked ensemble. The stacked ensemble parameters were: ensemble_level=2, transforming the 21 original features to 72 transformed features. Each of the three stacked models were fit on three internal holdout splits and were linearly combined. Similarly, the predictive model with top eleven features was a stacked ensemble. The stacked ensemble parameters were: ensemble_level=2, transforming the 11 original features to 34 transformed features. Each of the three stacked models were fit on three internal holdout splits and were linearly combined. The fitted features of the final model were the best features found during the feature engineering iterations.

**Supplemental Figure 3.**
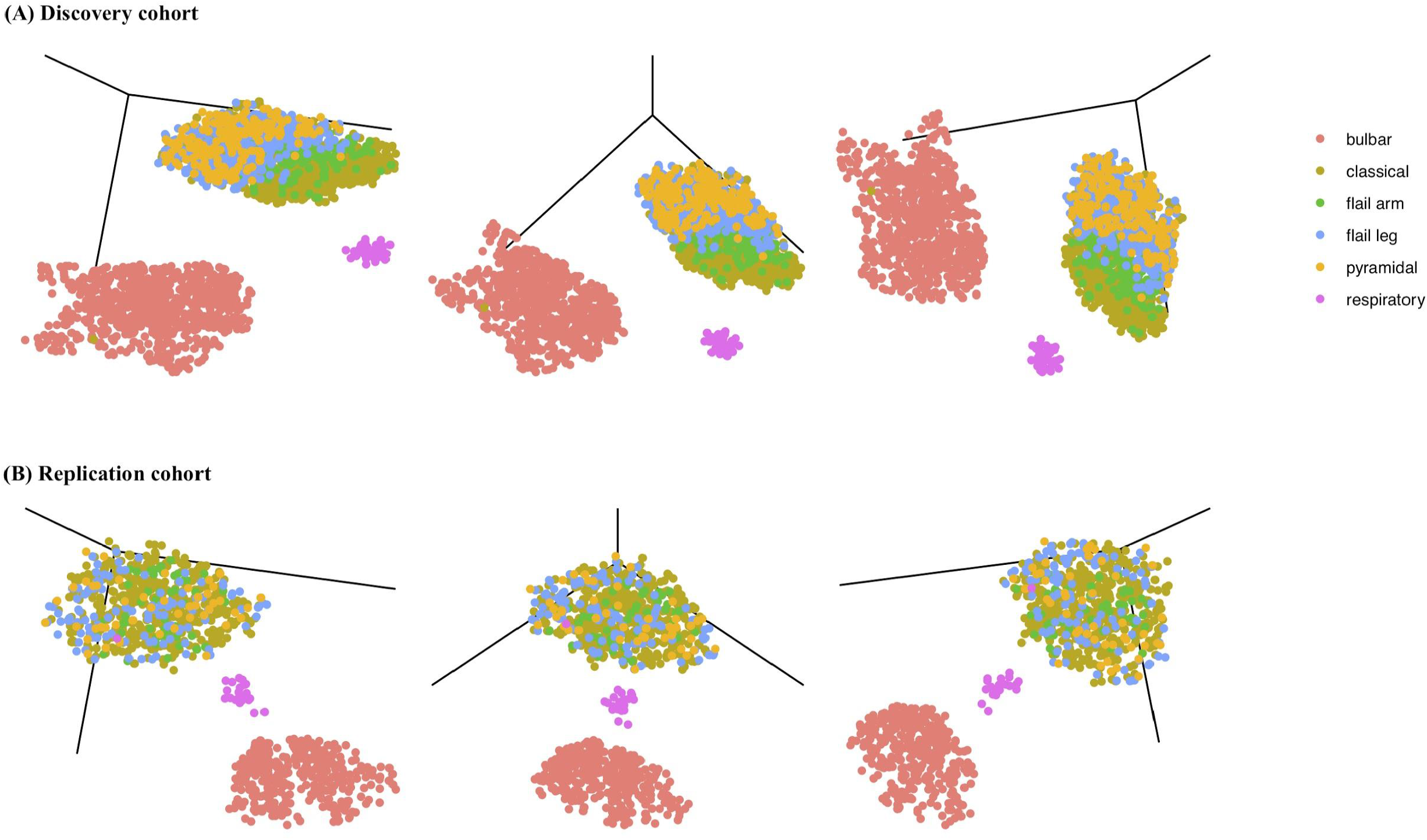
ALS subtypes identified by machine learning in the discovery and replication cohorts using UMAP alone. The top row (A) shows the three different 3D projections of the discovery cohort defined by the semi-supervised machine learning algorithm consisting of a UMAP algorithm alone. The same 3D projections of the replication cohort are shown in the bottom row (B). Each patient (dot) was color-coded after machine learning cluster generation according to the Chiò classification system.

**Supplemental Figure S4.**
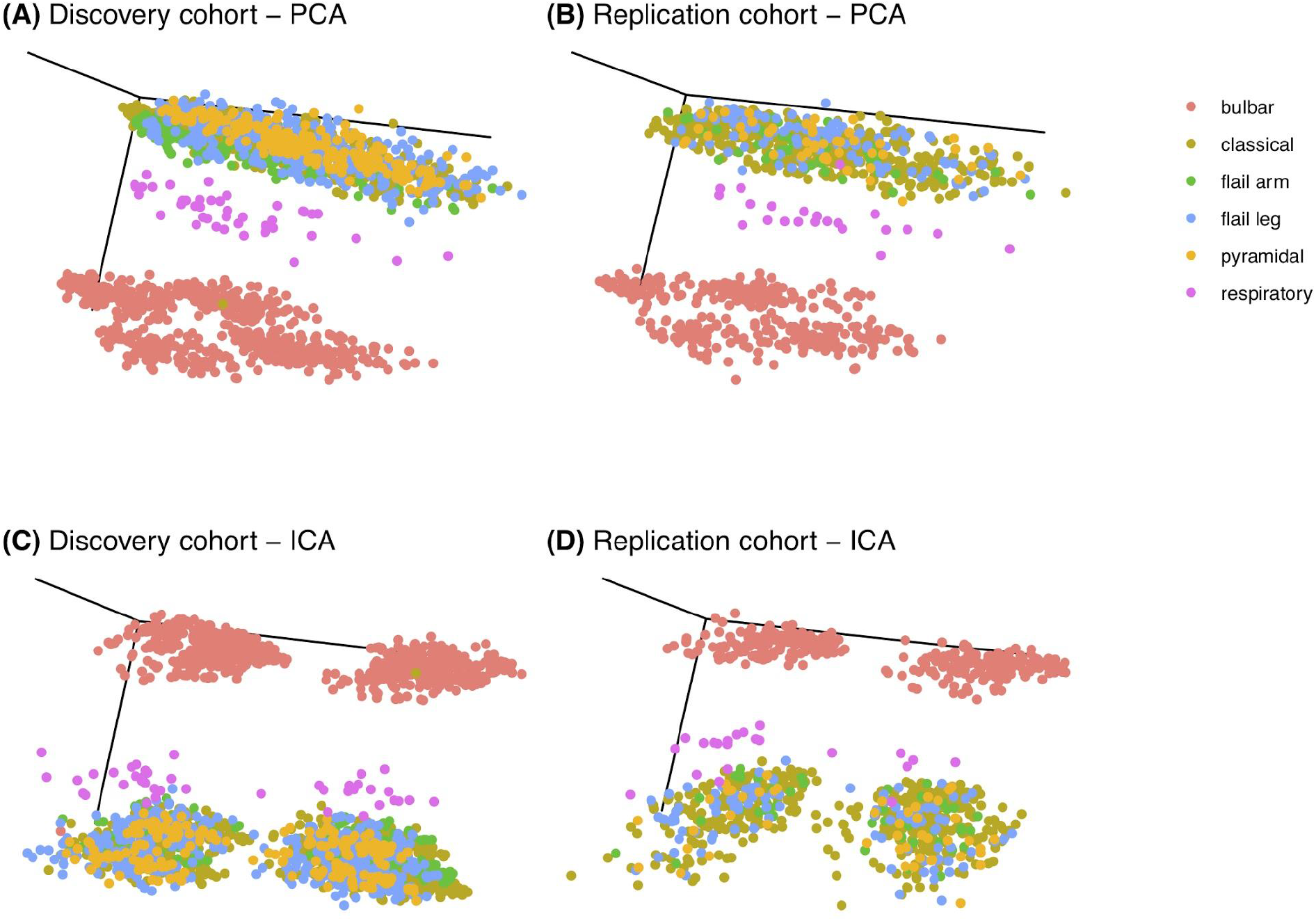
ALS clustering using dimension reduction approaches principal component analysis (PCA) and independent component analysis (ICA). The top row shows a 3D projection of the discovery cohort (A) and the replication cohort (B) clustered using PCA. The bottom row shows the 3D projection of the discovery cohort (C) and the replication cohort (D) clustered using ICA. Each patient (dot) was color-coded after machine learning cluster generation according to the Chiò classification system.

**Supplemental Figure S5.**
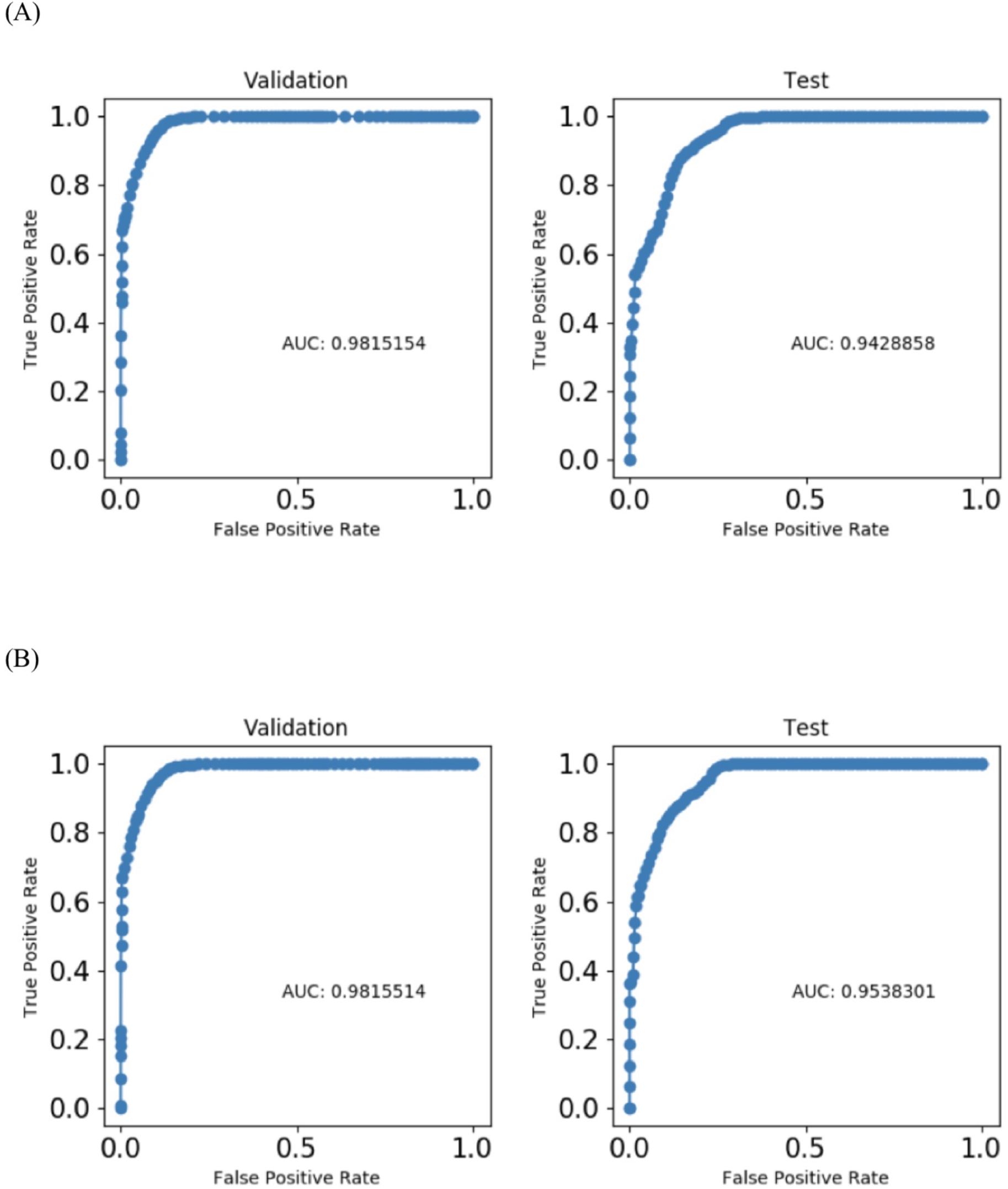
The performance of ALS disease subtype prediction models based on receive operating characteristic (ROC) curves. (A) ROC curves for the model that includes all features. The left panel shows the validation dataset and the right panel shows the test dataset. (B) ROC curves for the model based on the twelve most important features. The left panel shows the validation dataset and the right panel shows the test dataset.

**Supplemental Figure S6.**
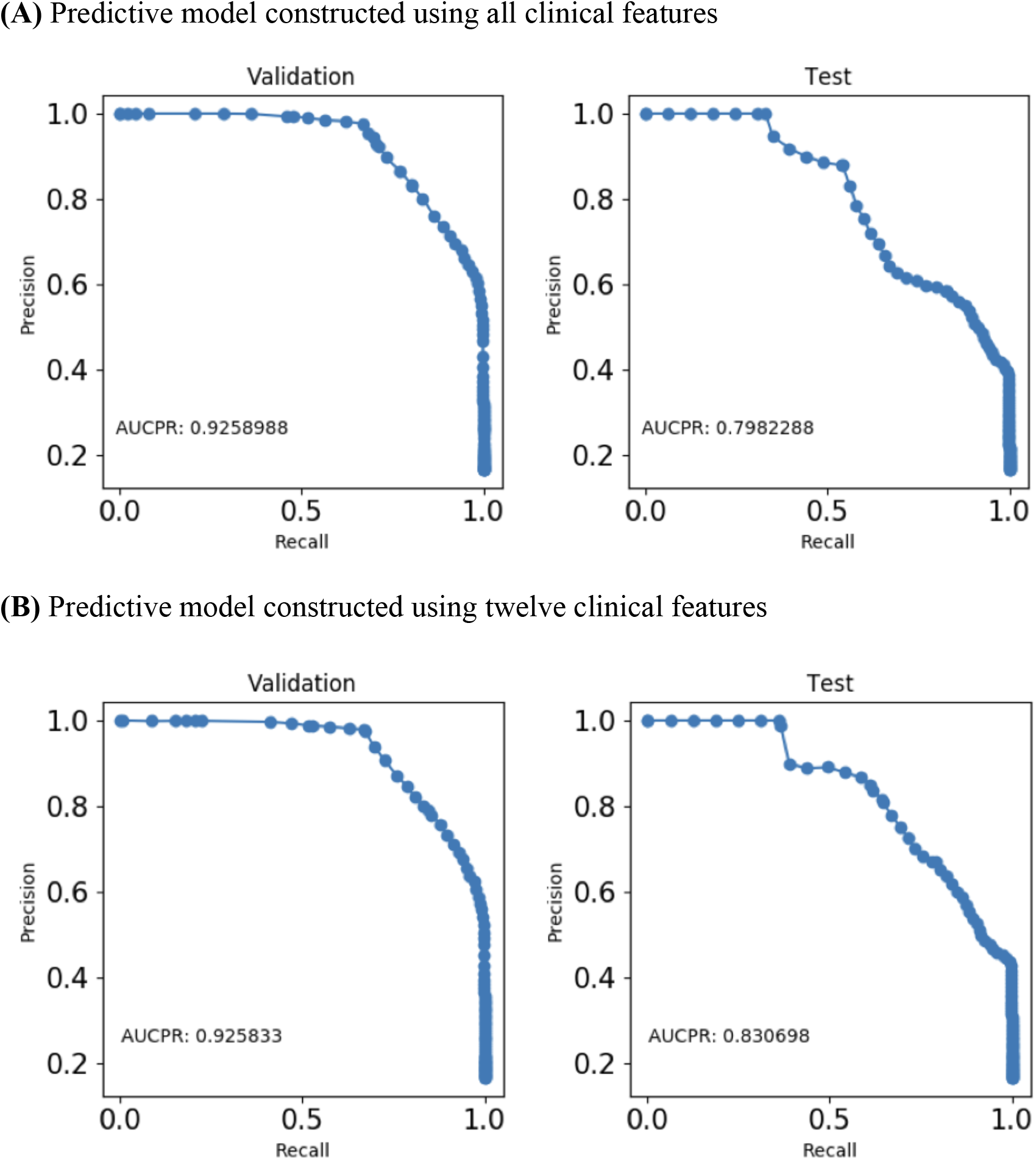
Precision Recall Curve for performance of ALS disease subtype prediction models. The top row (**A**) shows the precision recall curve for the prediction model based on all the clinical features. The bottom row (**B**) shows the precision recall curve for the prediction model based on the twelve most important features.

**Supplemental Table S1.**
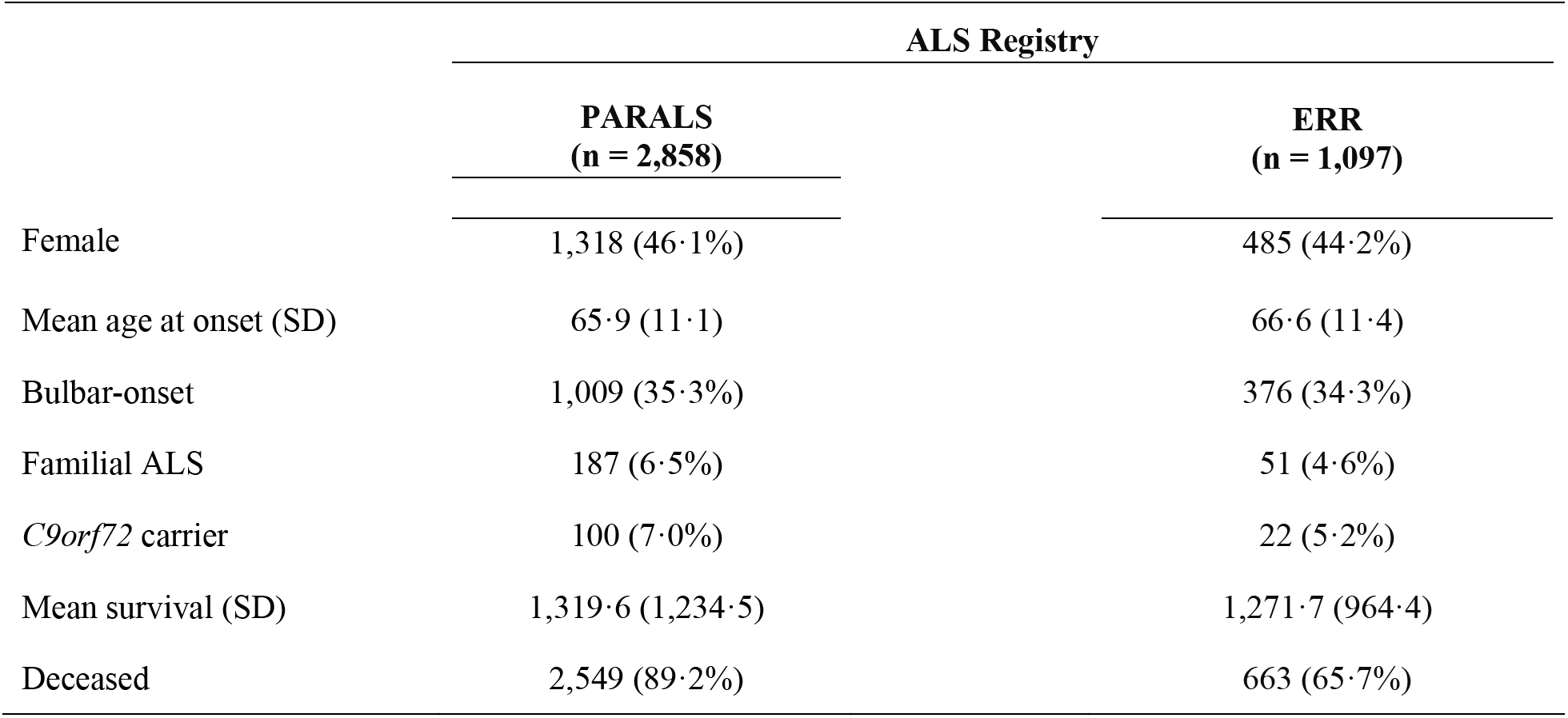
Demographics and clinical features of the ALS registries analyzed in this study.

**Supplemental Table S2.** Clinical parameters of the discovery and replication ALS cohorts used in each stage of analysis.

|  | Prediction models |  |
| --- | --- | --- |
|  | Unsupervised and semi-supervised | Supervised |
| Cancer | Yes | No |
| Cancer type | No (irrelevance) | No |
| Hypertension | Yes | No |
| Hyperthyroid | Yes | No |
| Hypothyroid | Yes | No |
| Diabetes | Yes | No |
| COPD | Yes | No |
| Smoker | No (missingness) | Yes |
| Parkinsonism | Yes | No |
| Chorea | Yes | No |
| Ataxia | Yes | No |
| Marital status | Yes | No |
| Education | Yes | No |
| Cognitive status 1 | No (missingness) | No |
| Cognitive status 2 | No (missingness) | No |
| Cognitive impairment present | No (missingness) | No |
| Initial diagnosis was PLS | No (bias) | No |
| El Escorial category at diagnosis | Yes | Yes |
| El Escorial category at visit 2 | No (irrelevance) | No |
| El Escorial category at visit 3 | No (irrelevance) | No |
| FVC percent at diagnosis | Yes | Yes |
| Height | Yes | No |
| Weight at diagnosis | Yes | Yes |
| BMI at diagnosis | Yes | No |
| Weight 2 years prior to illness | Yes | No |
| BMI 2 years prior to illness | Yes | No |
| Rate of decline BMI (per month) | Yes | Yes |
| PEG inserted | No (data leakage) | No |
| Time of PEG (days into illness) | No (data leakage) | No |
| NIPPV | No (data leakage) | No |
| Time of NIPPV (days into illness) | No (data leakage) | No |
| Tracheostomy | No (data leakage) | No |
| Time of tracheostomy (days into illness) | No (data leakage) | No |
| Place of birth | No | No |
| Place of residence | No | No |
| Family history of ALS | Yes | No |
| *C9orf72* status | No (data leakage) | No |
| Mutation present | No (bias) | No |
| Mutated gene | No (bias) | No |
| Mutation amino acid change | No (bias) | No |
| Anatomical level at onset | Yes | Yes |
| Site of symptom onset | Yes | Yes |
| Onset side | Yes | Yes |
| Clinical type at onset | No (missingness) | No |
| Clinical type at one year | No (data leakage) | No |
| Sex | Yes | No |
| Age at symptom onset | Yes | Yes |
| Age at diagnosis | Yes | No |
| Delay in diagnosis (days) | Yes | No |
| Survival (days) | No (data leakage) | No |
| Vital status | No (data leakage) | No |
| ALSFRS1 | Yes | Yes |
| ALSFRS2 | Yes | No |
| ALSFRS3 | Yes | No |
| ALSFRS4 | Yes | No |
| ALSFRS5 | Yes | No |
| ALSFRS6 | Yes | No |
| ALSFRS7 | Yes | No |
| ALSFRS8 | Yes | No |
| ALSFRS9 | Yes | No |
| ALSFRS10 | Yes | No |
| ALSFRS11 | Yes | No |
| ALSFRS12 | Yes | No |
| First ALSFRS-R total | Yes | No |
| Time of first ALSFRS-R (days into | Yes | Yes |
| Rate of decline ALSFRS-R (per month) | Yes | No |

ALSFRS-R, ALS Functional Rating Scale – Revised; ALSFRS1 refers to the first question on the scale. ALSFRS2, second question; BMI, body mass index; COPD, chronic obstructive pulmonary disease; PEG, percutaneous endoscopic gastrostomy; NIPPV, non-invasive positive pressure ventilation.

**Supplementary Table S3.** Performance of the models used to predict ALS clinical subtypes. The tables show the metrics used to evaluate the performance of (A) the model based on all of the available clinical features and (B) the model based on only the most important clinical features.

| Scorer | Optimized | Better score is | Final ensemble scores on validation (internal) | Final ensemble standard deviation on validation (internal) | Final test scores (external) | Final test standard deviation |
| --- | --- | --- | --- | --- | --- | --- |
| ACCURACY |  | higher | 0·8150798 | 0·01275987 | 0·6439888 | 0·01293791 |
| AUC |  | higher | 0·9815154 | 0·002010137 | 0·9428858 | 0·003738604 |
| AUCPR |  | higher | 0·9258988 | 0·00742477 | 0·7982288 | 0·01136424 |
| F05 |  | higher | 0·8150798 | 0·01275987 | 0·6439888 | 0·01293791 |
| F1 |  | higher | 0·8150798 | 0·01275987 | 0·6439888 | 0·01293791 |
| F2 |  | higher | 0·8150798 | 0·01275987 | 0·6439888 | 0·01293791 |
| GINI |  | higher | 0·9630309 | 0·004020274 | 0·8857717 | 0·007477209 |
| LOGLOSS | * | lower | 0·4072223 | 0·02122787 | 0·7757664 | 0·03125351 |
| MACROAUC |  | higher | 0·9585233 | 0·003836152 | 0·8972699 | 0·006460401 |
| MCC |  | higher | 0·7780957 | 0·01531184 | 0·5727866 | 0·01552549 |

(B) Predictive model based on most important clinical features
| Scorer | Optimized | Better score is | Final ensemble scores on validation (internal) | Final ensemble standard deviation on validation (internal) | Final test scores (external) | Final test standard deviation |
| --- | --- | --- | --- | --- | --- | --- |
| ACCURACY |  | higher | 0·8145004 | 0·009715746 | 0·7148183 | 0·01396052 |
| AUC |  | higher | 0·9815514 | 0·001332268 | 0·9538301 | 0·003653627 |
| AUCPR |  | higher | 0·925833 | 0·005046326 | 0·830698 | 0·01184665 |
| F05 |  | higher | 0·8145004 | 0·009715746 | 0·7148183 | 0·01396052 |
| F1 |  | higher | 0·8145004 | 0·009715746 | 0·7148183 | 0·01396052 |
| F2 |  | higher | 0·8145004 | 0·009715746 | 0·7148183 | 0·01396052 |
| GINI |  | higher | 0·9631028 | 0·002664535 | 0·9076601 | 0·007307254 |
| LOGLOSS | * | lower | 0·411088 | 0·0143602 | 0·7687869 | 0·03917954 |
| MACROAUC |  | higher | 0·9595809 | 0·002420295 | 0·9051199 | 0·006400558 |
| MCC |  | higher | 0·7774005 | 0·0116589 | 0·6577819 | 0·01675263 |

**Supplementary Table S4.** Validation Confusion Matrix of the models used to predict ALS clinical subtypes. The tables show the validation confusion matrix of (A) the model based on all of the available clinical features and (B) the model based on only the most important clinical features.

|  | Predicted:<br>bulbar | Predicted:<br>classical | Predicted:<br>flail arm | Predicted:<br>flail leg | Predicted:<br>pyramidal | Predicted:<br>respiratory | error |
| --- | --- | --- | --- | --- | --- | --- | --- |
| Actual: bulbar | 1,007 | 1 | 0 | 1 | 0 | 0 | 0% |
| Actual: classical | 1 | 601 | 1 | 223 | 15 | 0 | 29% |
| Actual: flail arm | 0 | 28 | 160 | 0 | 0 | 0 | 15% |
| Actual: flail leg | 0 | 34 | 0 | 470 | 27 | 0 | 11% |
| Actual: pyramidal | 0 | 11 | 0 | 189 | 40 | 0 | 83% |
| Actual: respiratory | 0 | 0 | 0 | 0 | 0 | 48 | 0% |

(B) Predictive model based on most important clinical features
|  | Predicted:<br>bulbar | Predicted:<br>classical | Predicted:<br>flail arm | Predicted:<br>flail leg | Predicted:<br>pyramidal | Predicted:<br>respiratory | error |
| --- | --- | --- | --- | --- | --- | --- | --- |
| Actual: bulbar | 1,007 | 1 | 0 | 1 | 0 | 0 | 0% |
| Actual: classical | 1 | 657 | 1 | 156 | 26 | 0 | 22% |
| Actual: flail arm | 0 | 28 | 160 | 0 | 0 | 0 | 15% |
| Actual: flail leg | 0 | 91 | 0 | 397 | 43 | 0 | 25% |
| Actual: pyramidal | 0 | 46 | 0 | 137 | 57 | 0 | 76% |
| Actual: respiratory | 0 | 0 | 0 | 0 | 0 | 48 | 0% |

**Supplementary Table S5.**
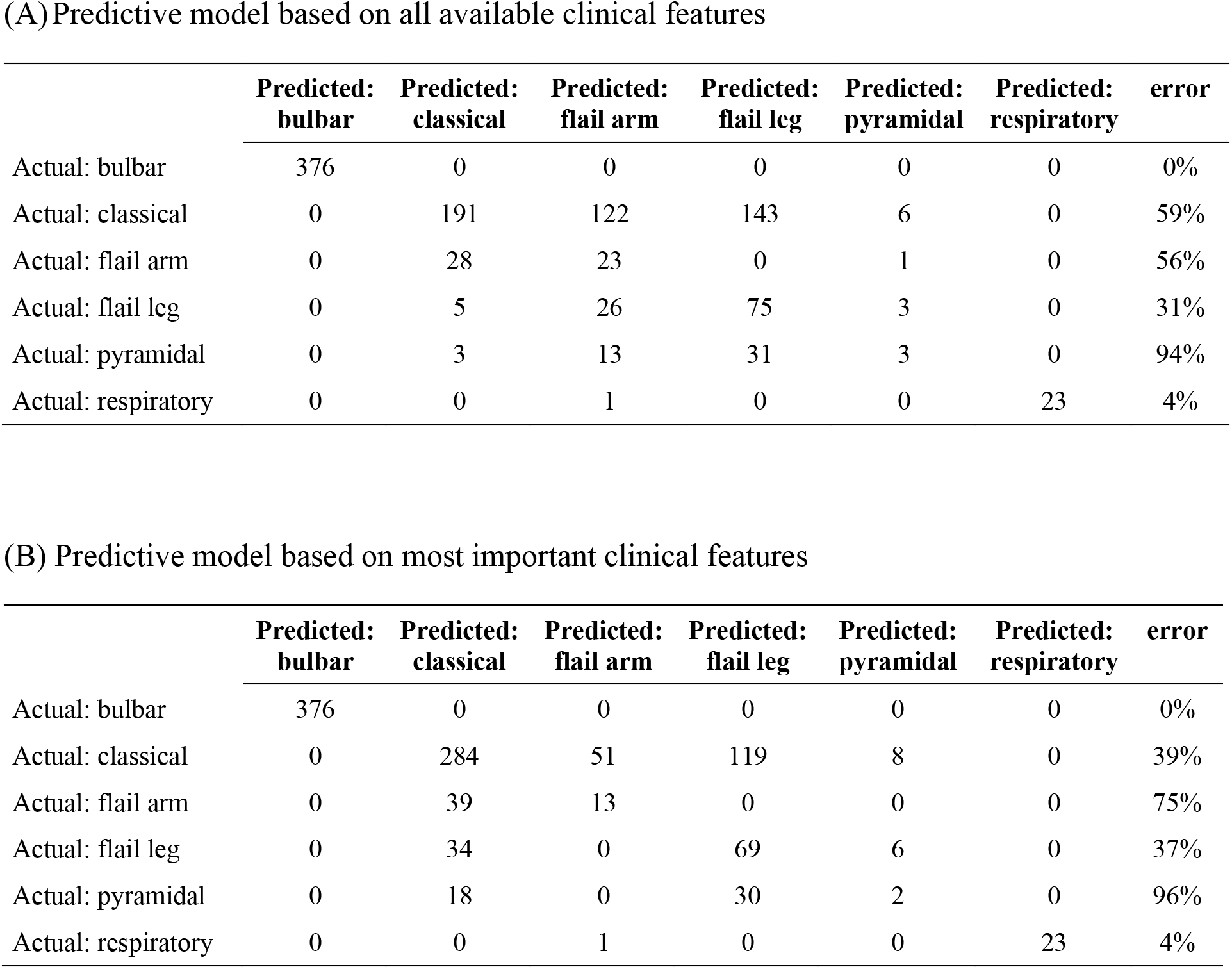
Test Confusion Matrix of the models used to predict ALS clinical subtypes. The tables show the test confusion matrix of (A) the model based on all of the available clinical features and (B) the model based on only the most important clinical features.

|  | Predicted:<br>bulbar | Predicted:<br>classical | Predicted:<br>flail arm | Predicted:<br>flail leg | Predicted:<br>pyramidal | Predicted:<br>respiratory | error |
| --- | --- | --- | --- | --- | --- | --- | --- |
| Actual: bulbar | 376 | 0 | 0 | 0 | 0 | 0 | 0% |
| Actual: classical | 0 | 191 | 122 | 143 | 6 | 0 | 59% |
| Actual: flail arm | 0 | 28 | 23 | 0 | 1 | 0 | 56% |
| Actual: flail leg | 0 | 5 | 26 | 75 | 3 | 0 | 31% |
| Actual: pyramidal | 0 | 3 | 13 | 31 | 3 | 0 | 94% |
| Actual: respiratory | 0 | 0 | 1 | 0 | 0 | 23 | 4% |

(B) Predictive model based on most important clinical features
|  | Predicted:<br>bulbar | Predicted:<br>classical | Predicted:<br>flail arm | Predicted:<br>flail leg | Predicted:<br>pyramidal | Predicted:<br>respiratory | error |
| --- | --- | --- | --- | --- | --- | --- | --- |
| Actual: bulbar | 376 | 0 | 0 | 0 | 0 | 0 | 0% |
| Actual: classical | 0 | 284 | 51 | 119 | 8 | 0 | 39% |
| Actual: flail arm | 0 | 39 | 13 | 0 | 0 | 0 | 75% |
| Actual: flail leg | 0 | 34 | 0 | 69 | 6 | 0 | 37% |
| Actual: pyramidal | 0 | 18 | 0 | 30 | 2 | 0 | 96% |
| Actual: respiratory | 0 | 0 | 1 | 0 | 0 | 23 | 4% |

## REFERENCES

1. Arthur KC, Calvo A, Price TR, Geiger JT, Chiò A, Traynor BJ. Projected increase in amyotrophic lateral sclerosis from 2015 to 2040. Nat Commun 2016; 7: 12408.

2. Hirtz D, Thurman DJ, Gwinn-Hardy K, Mohamed M, Chaudhuri AR, Zalutsky R. How common are the “common” neurologic disorders? Neurology 2007; 68(5): 326–37.

3. Chia R, Chiò A, Traynor BJ. Novel genes associated with amyotrophic lateral sclerosis: diagnostic and clinical implications. Lancet Neurol 2018; 17(1): 94–102.

4. Turner MR, Hardiman O, Benatar M, et al. Controversies and priorities in amyotrophic lateral sclerosis. Lancet Neurol 2013; 12(3): 310–22.

5. Byrne S, Bede P, Elamin M, et al. Proposed criteria for familial amyotrophic lateral sclerosis. Amyotroph Lateral Scler 2011; 12(3): 157–9.

6. Roche JC, Rojas-Garcia R, Scott KM, et al. A proposed staging system for amyotrophic lateral sclerosis. Brain 2012; 135(Pt 3): 847–52.

7. de Carvalho M, Dengler R, Eisen A, et al. Electrodiagnostic criteria for diagnosis of ALS. Clin Neurophysiol 2008; 119(3): 497–503.

8. Geevasinga N, Howells J, Menon P, et al. Amyotrophic lateral sclerosis diagnostic index: Toward a personalized diagnosis of ALS. Neurology 2019; 92(6): e536–e47.

9. Brooks BR. El Escorial World Federation of Neurology criteria for the diagnosis of amyotrophic lateral sclerosis. Subcommittee on Motor Neuron Diseases/Amyotrophic Lateral Sclerosis of the World Federation of Neurology Research Group on Neuromuscular Diseases and the El Escorial “Clinical limits of amyotrophic lateral sclerosis” workshop contributors. J Neurol Sci 1994; 124 **Suppl**: 96-107.

10. Piemonte, Valle d’Aosta Register for Amyotrophic Lateral S. Incidence of ALS in Italy: evidence for a uniform frequency in Western countries. Neurology 2001; 56(2): 239–44.

11. Mandrioli J, Biguzzi S, Guidi C, et al. Epidemiology of amyotrophic lateral sclerosis in Emilia Romagna Region (Italy): A population based study. Amyotroph Lateral Scler Frontotemporal Degener 2014; 15(3-4): 262–8.

12. Cedarbaum JM, Stambler N, Malta E, et al. The ALSFRS-R: a revised ALS functional rating scale that incorporates assessments of respiratory function. BDNF ALS Study Group (Phase III). J Neurol Sci 1999; 169(1-2): 13–21.

13. Beretta L, Santaniello A. Nearest neighbor imputation algorithms: a critical evaluation. BMC Med Inform Decis Mak 2016; 16 Suppl 3: 74.

14. Orhan U, Hekim M, Ozer M. EEG signals classification using the K-means clustering and a multilayer perceptron neural network model. Expert Systems with Applications 2011; 38: 13475–81.

15. McInnes L, Healy J, Saul N, Großberger L. UMAP: Uniform Manifold Approximation and Projection. Journal of Open Source Software 2018; 3(29): 861.

16. Rokach L. Ensemble-based classifiers. Artificial Intelligence Review 2010; 33(1): 1–39.

17. Breiman L. Random Forests. Machine Learning 2001; 45(1): 5–32.

18. Ke G, Meng Q, Finley T. LightGBM: A Highly Efficient Gradient Boosting Decision Tree. In: I. G, UV. L, S. B, eds. Advances in Neural Information Processing Systems: Curran Associates, Inc.; 2017: 3146-54.

19. Chen T, Guestrin C. XGBoost: A Scalable Tree Boosting System. Proceedings of the 22nd ACM SIGKDD International Conference on Knowledge Discovery and Data Mining. New York: ACM; 2016: 785-94.

20. Lundberg S, Lee S. A unified approach to interpreting model predictions. In: I. G, U.V. L, S. B, et al., editors. 31st Conference on Neural Information Processing Systems (NIPS 2017); Long Beach, CA, USA: Curran Associates, Inc. p. 4765-74.

21. Maddox TM, Rumsfeld JS, Payne PRO. Questions for Artificial Intelligence in Health Care. JAMA 2019; 321(1): 31–2.

22. Verghese A, Shah NH, Harrington RA. What This Computer Needs Is a Physician: Humanism and Artificial Intelligence. JAMA 2018; 319(1): 19–20.

23. Chiò A, Calvo A, Moglia C, Mazzini L, Mora G, group Ps. Phenotypic heterogeneity of amyotrophic lateral sclerosis: a population based study. J Neurol Neurosurg Psychiatry 2011; 82(7): 740–6.

24. Chiò A, Hammond ER, Mora G, Bonito V, Filippini G. Development and evaluation of a clinical staging system for amyotrophic lateral sclerosis. J Neurol Neurosurg Psychiatry 2015; 86(1): 38–44.

25. Kueffner R, Zach N, Bronfeld M, et al. Stratification of amyotrophic lateral sclerosis patients: a crowdsourcing approach. Sci Rep 2019; 9(1): 690.

26. Kuffner R, Zach N, Norel R, et al. Crowdsourced analysis of clinical trial data to predict amyotrophic lateral sclerosis progression. Nat Biotechnol 2015; 33(1): 51–7.

27. Leonard H, Blauwendraat C, Krohn L, et al. Genetic variability and potential effects on clinical trial outcomes: perspectives in Parkinson’s disease. J Med Genet 2020; 57(5): 331–8.

28. Stevens LM, Mortazavi BJ, Deo RC, Curtis L, Kao DP. Recommendations for Reporting Machine Learning Analyses in Clinical Research. Circ Cardiovasc Qual Outcomes 2020; 13(10): e006556.

29. Atassi N, Berry J, Shui A, et al. The PRO-ACT database: design, initial analyses, and predictive features. Neurology 2014; 83(19): 1719–25.

